# Early drivers of clonal hematopoiesis shape the evolutionary trajectories of *de novo* acute myeloid leukemia

**DOI:** 10.1101/2024.08.31.24312756

**Authors:** Ryan D. Chow, Priya Velu, Safoora Deihimi, Jonathan Belman, Angela Youn, Nisargbhai Shah, Selina M. Luger, Martin P. Carroll, Jennifer Morrissette, Robert L Bowman

## Abstract

Mutations commonly found in AML such as *DNMT3A*, *TET2* and *ASXL1* can be found in the peripheral blood of otherwise healthy adults – a phenomenon referred to as clonal hematopoiesis (CH). These mutations are thought to represent the earliest genetic events in the evolution of AML. Genomic studies on samples acquired at diagnosis, remission, and at relapse have demonstrated significant stability of CH mutations following induction chemotherapy. Meanwhile, later mutations in genes such as *NPM1* and *FLT3*, have been shown to contract at remission and in the case of *FLT3* often are absent at relapse. We sought to understand how early CH mutations influence subsequent evolutionary trajectories throughout remission and relapse in response to induction chemotherapy. Here, we assembled a retrospective cohort of patients diagnosed with *de novo* AML at our institution that underwent genomic sequencing at diagnosis as well as at the time of remission and/or relapse (total n = 182 patients). Corroborating prior studies, *FLT3* and *NPM1* mutations were generally eliminated at the time of cytologic complete remission but subsequently reemerged upon relapse, whereas *DNMT3A*, *TET2* and *ASXL1* mutations often persisted through remission. Early CH-related mutations exhibited distinct constellations of co-occurring genetic alterations, with *NPM1* and *FLT3* mutations enriched in *DNMT3A*^mut^ AML, while *CBL* and *SRSF2* mutations were enriched in *TET2*^mut^ and *ASXL1^mut^* AML, respectively. In the case of *NPM1* and *FLT3* mutations, these differences vanished at the time of complete remission yet readily reemerged upon relapse, indicating the reproducible nature of these genetic interactions. Thus, early CH-associated mutations that precede malignant transformation subsequently shape the evolutionary trajectories of AML through diagnosis, therapy, and relapse.

**Key Points:**

- *DNMT3A*, *TET2* and *ASXL1* mutations persist through AML-directed therapy
- Distinct CH-related mutations shape the evolutionary trajectories of AML from diagnosis through relapse.

## INTRODUCTION

Acute myeloid leukemia (AML) results from the accumulation of genetic alterations in hematopoietic stem/progenitor cells, leading to clonal expansion and impaired differentiation.^1^ While extensive work has been devoted to profiling the genomic aberrations that define AML^2–6^, many common AML-associated mutations are also detected with increasing age in patients with otherwise intact hematopoietic function.^7–12^ The acquisition of somatic mutations that result in clonal expansion – termed clonal hematopoiesis (CH) – is associated with increased risk of developing not only hematologic malignancies, but also a host of other diseases.^7,8^ While only a fraction of patients with CH will ultimately be diagnosed with a hematologic malignancy, these somatic mutant clones are nevertheless thought to represent preleukemic precursors that are primed for malignant transformation upon the acquisition of further driver mutations.^13,14^ Of note, CH-associated mutations can be detected in the peripheral blood of patients that have achieved complete remission from AML and have been identified in hematopoietic stem/progenitor cells that survive chemotherapy.^15–19^ Thus, the mutational drivers of CH likely represent the earliest genetic events in the pathogenesis of AML, providing the substrate for further genomic evolution and malignant transformation.

Extensive genomic profiling has been performed in *de novo* and secondary AML, highlighting trends of genomic evolution for *FLT3* or *NPM1* mutant disease.^20–23^ Additionally, other studies have described paired longitudinal sequencing of patient samples undergoing *FLT3* tyrosine kinase inhibition treatment^24,25^ or following induction chemotherapy.^26,27^ More recently, the advent of single cell DNA sequencing^28–30^ and error corrected sequencing^31–33^ has dramatically improved the evaluation of mutation evolution during remission, highlighting both the stability of clonal hematopoiesis mutations following therapy^34,35^ and the need to eradicate even the smallest of *FLT3* and *NPM1* mutant clones to control disease progression.^36,37^ To date, however, few cohorts have evaluated serial samples of patients from diagnosis through remission and subsequent relapse, connecting genomic trajectories through several stages of disease management.

Here, we assembled a cohort of patients diagnosed with *de novo* AML at our institution between 2013-2018. To investigate clonal evolution throughout diagnosis and disease management, we selected patients that underwent next-generation sequencing (NGS) both at diagnosis and again at remission and/or relapse. We find that *DNMT3A*^mut^, *TET2*^mut^, and *ASXL1*^mut^ (DTA) AML exhibit distinct mutational profiles at diagnosis, and that these differences persist through the selective pressure of chemotherapy. Thus, we demonstrate that early preleukemic drivers of CH can influence the subsequent evolutionary trajectories of AML.

## RESULTS

### Charting the genomic evolution of de novo AML at diagnosis, remission and relapse

We retrospectively compiled all patients diagnosed with *de novo* AML at our institution that had two or more NGS studies, performed at least 30 days apart, on blood or bone marrow specimens. The final cohort comprised 182 patients. The average age at diagnosis was 58.06 ± 1.03 (mean ± s.e.m.) years, and 53.3% of patients were female. In total, 84.6% of patients received induction chemotherapy with combination anthracycline and nucleoside analog therapy (i.e. “7+3”) (**Supplementary Figure 1A**), and 41.8% (n = 76) underwent stem cell transplant at some point in their treatment.

At the time of AML diagnosis, *FLT3* (38%), *NPM1* (32%), *DNMT3A* (32%), and *TET2* (21%) were the most frequently mutated genes in our cohort (**Figure 1A**). The mutation frequencies of the top mutated genes (defined as mutated in ≥ 4% of diagnosis samples) were highly correlated with those observed in the TCGA^4^ and BeatAML^38^ cohorts (**Supplementary Figure 1B-C**). Based on karyotype analysis, chromosome 8 gain (10%) and chromosome 16 inversion (5%) were the most frequent chromosomal abnormalities. Approximately half of the patient cohort underwent AML NGS profiling twice, while the remaining half had three or more NGS profiles (**Figure 1B**). In total, 65.3% of patients (n = 119) were sequenced at the time of first cytologic complete remission (CR1) and 41.8% (n = 76) at the time of first relapse (REL1) (**Figure 1C**).

**Figure 1:**
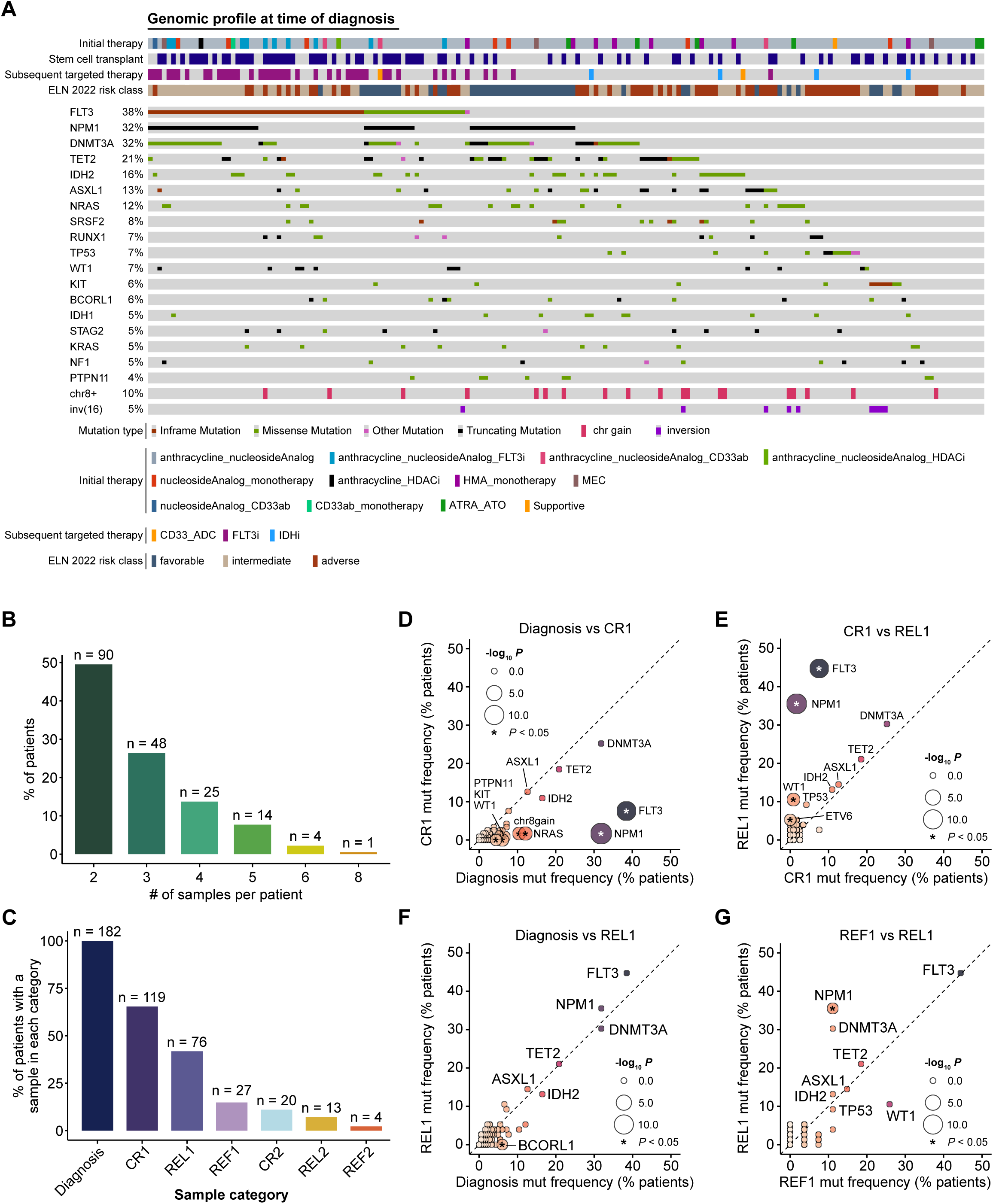
Charting the genomic evolution of *de novo* AML at diagnosis, remission and relapse. **A.** Frequently mutated genes and karyotype aberrations at time of diagnosis in the Penn AML cohort (total n = 182 patients). Patients are annotated by the treatments received throughout the course of the disease and by ELN 2022 risk classifications. **B.** Distribution of the number of serial genomic profiles obtained for each patient, expressed as a percentage of the total cohort. All patients included in the cohort underwent genomic profiling at least twice, with more than half having 3 or more matched genomic samples. The number of patients in each category is annotated above. **C.** Distribution of the number of patients with a genomic profile at each stage of AML disease progression, expressed as a percentage of the total cohort. The number of patients represented in each category is annotated above. CR1: first complete remission. REL1: first relapse. REF1: first refractory disease. CR2: second complete remission. REL2: second relapse. REF2: second refractory disease. **D-G**. Comparison of cohort-level mutation frequencies across different disease timepoints. (**D**) diagnosis (n = 182) vs CR1 (n = 119); (**E**) CR1 vs REL1 (n = 76); (**F**) diagnosis vs REL1; (**G**) REF1 (n = 27) vs REL1. Point sizes are scaled by statistical significance (Fisher’s two-sided exact test) and colored based on mutation frequency. Asterisks indicate *P* < 0.05. Dashed lines denote equality between disease stages. CR1, first remission; REL1, first relapse; REF1, first refractory disease; HDACi, histone deacetylase inhibitor.

To investigate how AML-directed therapy would affect the mutational landscape, we compared gene mutation frequencies across varying disease stages. Comparing samples taken at CR1 to those at diagnosis, we observed significant depletion of *FLT3*, *NPM1*, and *NRAS* mutations, as well as chromosome 8 copy gain (**Figure 1D**). In contrast, the CH-associated genes *DNMT3A*, *TET2*, and *ASXL1* were mutated at nearly identical frequencies between diagnosis and CR1, consistent with prior reports.^6,15–17^ However, when these patients subsequently relapsed after an initial remission, the mutation frequencies of *FLT3* and *NPM1* largely returned to pre-treatment baseline levels (**Figure 1E-F**). Comparison of REL1 and primary refractory (REF1) samples showed similar mutation frequencies across genes with the exception of *NPM1*, which was comparatively enriched in REL1 samples (**Figure 1G**). Collectively, these analyses illustrate the rise and fall of key driver mutations over the course of AML progression, highlighting the remarkable consistency in driver mutation frequencies between diagnosis and relapse despite the interceding selective pressure of chemotherapy.

### DTA mutations persist at the time of complete remission

We next investigated mutation persistence at CR1. We grouped genes into functional biological categories and evaluated the frequency at which each mutation was detected in paired diagnosis and relapse samples (**Figure 2A**). We observed robust persistence of mutations in genes related to DNA damage (10/15, 66.7%), CH-associated DTAI factors (*DNMT3A*, *TET2*, *ASXL1, IDH1* and *IDH2* combined: 83/130, 63.8%), and splicing (12/19, 63.2%), with less mutational persistence observed in genes associated with the Polycomb repressive complex (PRC; 11/28, 39.3%) and cohesin complex (4/16, 25%). Breaking these categories down into individual genes, we observed robust persistence of *IDH2* mutations in over half of cases (13/21, 61.9%), whereas a smaller fraction of *IDH1* mutations persisted at remission (2/8, 25%) (**Figure 2B**). In line with this, the variant allele frequencies (VAFs) for the two persistent *IDH1* mutations were 6.9% and 10.3%, compared to a mean VAF of 30.3% ± 5.1% (s.e.m.) for *IDH2* (**Figure 2C-D**). Similar to *IDH2*, the mean VAFs for *DNMT3A, TET2,* and *ASXL1* remained high at the time of remission (33.7% ± 3.6%, 33.8% ± 2.8%, 34.7% ± 5.0%, respectively), with most patients retaining these DTA mutations, often showing few differences between diagnosis and CR1 (**Figure 2E**). In addition, we found that variants in *RUNX1*, *SRSF2,* and *TP53* were also frequently identified at CR1 (**Supplemental Figure 2A**).

**Figure 2:**
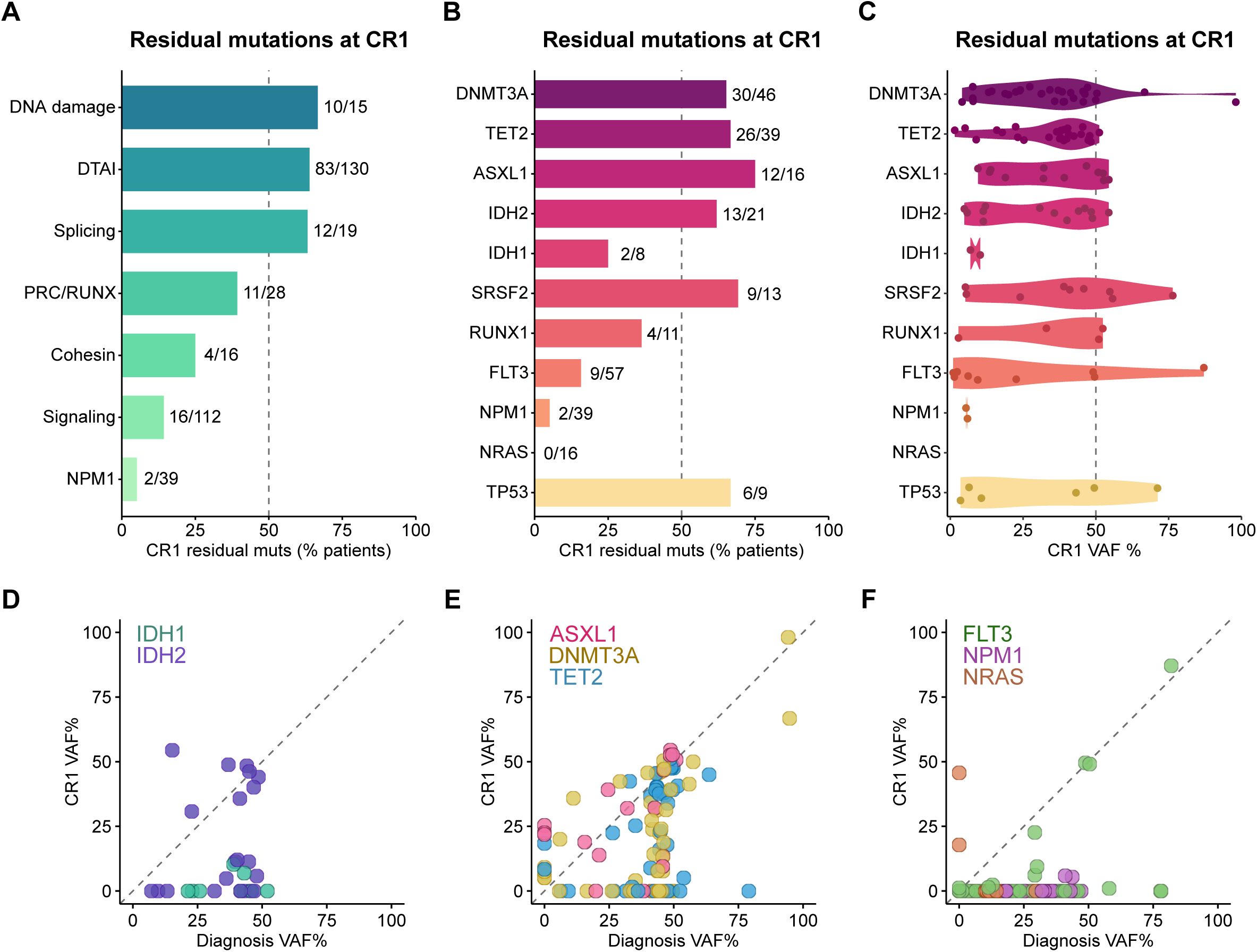
Mutations associated with clonal hematopoiesis are persistent at remission. **A.** Bar plot depicting the percentage of mutations identified in diagnosis that were also identified at first remission (CR1). Numbers to the right each bar indicate the proportion of variants initially found at diagnosis that were subsequently detected at CR1. Genes are grouped into their biological categories as relevant (DNA damage: *TP53*, *ATM*; DTAI: *DNMT3A*, *TET2*, *ASXL1*, *IDH2*, *IDH1*; Splicing: *SRSF2*, *U2AF1*, *ZRSR2*; PRC/RUNX: *BCOR*, *BCORL1*, *RUNX1*, *EZH2*; Cohesin: *SMC1A*, *RAD21*, *STAG2*; Signaling: *CSF1R*, *FLT3*, *NF1*, *KRAS*, *NRAS*, *BRAF*, *KIT*, *PTPN11*, *JAK2*, *CSF3R*, *CBL*). **B.** As in A, but individual genes are shown. **C.** Violin plot of VAFs for persistent variants at CR1. **D-F.** Scatterplot detailing patient-matched VAFs at diagnosis (x-axis) and CR1 (y-axis) for (**D**) *IDH1* and *IDH2*, (**E**) *DNMT3A, TET2,* and *ASXL1,* (**F**) *FLT3, NPM1,* and *NRAS*. Each point represents one variant in a specific patient, matched across time.

Finally, mutations in signaling genes including *FLT3, NRAS, KRAS,* and *PTPN11* were rarely maintained at CR1 (16/112, 14.3%), and we observed two instances of an *NPM1* mutation persisting at CR1 (2/39, 5.1%) (**Figure 2A)**. While we did not observe a single case of persistent *NRAS* mutations at CR1 (0/16), *FLT3* variants were shared between diagnosis and CR1 in 9/57 patients (16%); of these, four were at similar VAFs between diagnosis and CR1 (**Figure 2B,F**). Two of these were *FLT3* internal tandem duplications (ITDs) (E604-F605ins[11aa] and T582_E598dup), representing *bona fide* pathogenic mutations, while the remaining two variants were missense mutations of uncertain significance (V214I and V795I). In a similar manner, both of the identified persistent *NPM1* mutations were pathogenic W288Cfs*12 variants. Surprisingly, we identified one *FLT3* variant (Y572ins?) that was newly identified at CR1 compared to diagnosis (**Supplementary Figure 2B**). As this CR1-only *FLT3* variant was found at 1.24% VAF, it is likely that this variant was also present at diagnosis but had fallen short of the NGS detection and/or reporting threshold. We further observed rare persistent or acquired variants in genes such as *JAK2*, *TP53*, *U2AF1*, *NF1*, *SRSF2*, *ZRSR2*, *STAG2*, and *CBL* at the time of remission. (**Supplemental Figure 2B-C**).

Collectively, we observed that *DNMT3A*, *TET2,* and *ASXL1* variants were less likely to be eliminated by chemotherapy than *FLT3* or *NPM1* (DTA combined vs *FLT3*, *p* = 9.3 *10^-11^; DTA combined vs *NPM1*, *p* = 1.7 *10^-12^). Overall, these data indicate that CH-associated mutations frequently persist through chemotherapy at the time of remission in *de novo* AML, presumably due to their presence in a preleukemic cell compartment that remains intact despite effective AML-directed therapy.

### FLT3 variants are dynamically acquired and eliminated between diagnosis and relapse, while NPM1 mutations persist

We next compared the presence of matched variants at diagnosis compared to relapse (**Figure 3A**). We observed that all *ASXL1* variants (11/11, 100%) identified at diagnosis were also present at relapse. Similar results were evident with *DNMT3A* (28/31, 90.3%), *TET2* (25/26, 96.2%), *IDH2* (8/10, 80%) and *IDH1* (3/3, 100%). *NPM1* variants were similarly stable, with 27/32 (84.4%) shared between diagnosis and relapse. In comparing differences in VAFs between diagnosis and relapse, we observed that most of the DTAI mutations reemerged to a similar VAF compared to the time of diagnosis, indicating their likely presence in the initiating clone (*DNMT3A* 34.6% ± 3.0%, *TET2* 35.8% ±2.8%, *ASXL1* 45.2% ± 2.7%, *IDH1* 29.5% ±10.6%, *IDH2* 31.4% ± 4.9%) (**Figure 3B-D**). Similar results were evident for *NPM1* (**Figure 3E**). We did not observe an instance of a new mutation in *DNMT3A*, *TET2*, or *ASXL1* at time of first relapse, while we did observe gain of *IDH1* and *IDH2* mutations among our cohort. These comprised an *IDH1*^R132H^ mutation in a patient who was otherwise not mutated for any other DTAI or epigenetic factors represented in our NGS panel (**Supplemental Figure 3A**). In this patient, the emergent *IDH1*^R132H^ continued to expand through the subsequent line of therapy, further expanding to dominate the clonal composition. *IDH2*^R140Q^ mutations emerged at first relapse in two patients within the cohort, both of whom had co-occurring *DNMT3A*, *FLT3* and *NPM1* mutations at the time of diagnosis.

**Figure 3:**
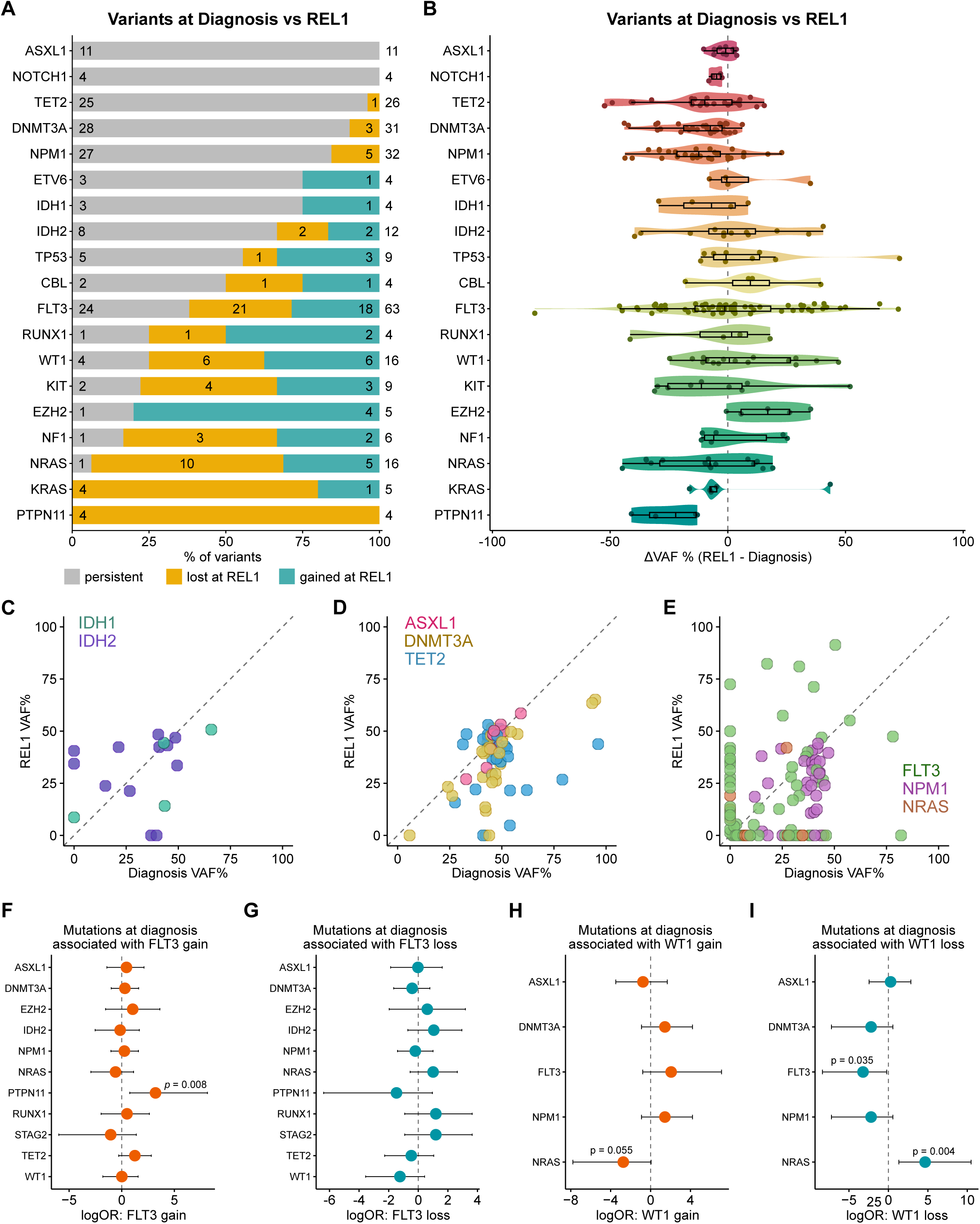
Signaling mutations undergo dynamic losses and gains from diagnosis through relapse. **A.** Bar plot depicting the relative proportions of different mutation trajectories between diagnosis and first relapse (REL1) in individual patients, filtered for genes with at least 4 variants identified across the cohort. Colors indicate whether the mutation was stable (grey), lost (yellow) or gained (teal) from diagnosis through relapse. The number to the right indicates the total number of variants identified among paired diagnosis and relapse samples for the indicated gene; within each section of the bar plot, the numbers indicate the number of variants within each category. **B.** Violin plot depicting the difference in VAFs (ΔVAF) between relapse and diagnosis. Negative values indicate a lower VAF at relapse, while positive values indicate a higher VAF. **C-E.** Scatterplot indicating VAF at diagnosis (x-axis) and REL1 (y-axis) for (**C**) *IDH1* and *IDH2*, (**D**) *DNMT3A, TET2,* and *ASXL1,* (**E**) *FLT3, NPM1,* and *NRAS*. Each point represents one variant in a specific patient, matched across time. **F-I.** Forest plot from Firth’s penalized logistic regression models evaluating the association between mutations at diagnosis in the indicated genes on the y-axis and (**F**) *FLT3* mutation gain, (**G**) *FLT3* mutation loss, (**H**) *WT1* mutation gain, and (**I**) *WT1* mutation loss. Points indicate the log odds ratios (logORs), with 95% confidence intervals.

In contrast to the CH-associated DTAI genes, mutations in signaling genes were largely unstable, with many being lost between diagnosis and relapse. These included *PTPN11* (4/4), *NRAS* (10/11, 90.9%)*, KRAS* (4/4), and *NF1* (3/4, 75%) (**Figure 3A**). *FLT3* mutations showed a more dynamic pattern compared to the other signaling mutations, with 53.3% (24/45) persisting from diagnosis to relapse. Of all identified *FLT3* variants in patients with paired diagnosis and REL1 samples, 18/63 (29%) were newly acquired upon relapse. We observed additional mutations with evidence of dynamic gains/losses including *EZH2, NRAS, RUNX1, TP53* and *KIT* (**Figure 3A**). Mutations in *WT1* were particularly dynamic, with six lost variants at REL1, six acquired variants, and four stable patients, mirroring the diverse range of outcomes present in *FLT3-*mutant disease. To identify co-mutational partners at diagnosis that might predict this evolution, we constructed Firth’s penalized regression models to determine the association between mutations at diagnosis and subsequent gain or loss of *FLT3* or *WT1* mutations. We identified a significant increase in the likelihood of gaining a *FLT3* mutation at relapse for patients that presented with a *PTPN11* mutation at diagnosis (*p=*0.008; **Figure 3F**); meanwhile no significant associations were identified that were associated a loss of *FLT3* mutation at relapse (**Figure 3G**). In one example, Patient 100 initially was found to have *IDH1*, *DNMT3A*, *NPM1*, and *PTPN11* mutations on diagnosis; at the time of relapse, the dominant population in this patient had lost the *PTPN11* mutation and instead gained a *FLT3* mutation (**Supplementary Figure 3B**). Similarly, Patient 156 initially had *DNMT3A* and *PTPN11* mutations, the latter of which was eliminated by chemotherapy and undetectable at CR1; at the time of REL1, the *PTPN11* mutation had been replaced by a *FLT3* mutation (**Supplementary Figure 3C**). Meanwhile, loss of *WT1* mutations were associated with *FLT3* and *NRAS* mutations, but in opposing directions **Figure 3H,I**). The presence of a *FLT3* mutation at diagnosis was associated with lower probability of *WT1* mutation loss (*p=*0.035), whereas the presence of an *NRAS* mutations was associated with in increased likelihood for loss of *WT1* mutations (*p=*0.004). Collectively, these results indicated that co-mutational partners are associated with distinct evolutionary outcomes following induction chemotherapy. While *FLT3* mutations are dynamically acquired and eliminated between diagnosis and relapse, CH-related mutations and *NPM1* mutations largely persist through complete remission into relapse.

### Co-mutation analyses reveal conserved and disease stage-specific genetic interactions in AML

To further explore this concept of dynamic co-mutational partners we identified co-occurring and mutually exclusive mutation pairs at the time of diagnosis (**Supplementary Figure 4A**), CR1 (**Supplementary Figure 4B**), and REL1 **(Supplementary Figure 4C**). We identified eight putative genetic interactions that were conserved between diagnosis and CR1: co-occurrence of *ASXL1*-*STAG2*, *ASXL1-SRSF2*, *BCOR1-SF1*, *BIRC3-MYD88*, *CBL-TET2*, *CDKN2A-PRPF40B*, chr5qdel-*TP53*, and *IDH2-SRSF2*. We further identified ten putative genetic interactions that were shared between diagnosis and REL1: co-occurrence of *CBL-TET2*, *CDH2*-t(8;21), *CDH2-CSF3R*, chr17loss-*TP53*, *DNMT3A-NPM1*, *DNMT3A-FLT3*, *FLT3-NPM1*, *NOTCH2-U2AF2*, *NPM1-TET2*, along with mutual exclusivity of *NPM1-TP53*. Co-occurrence of the *CBL-TET2* mutation pair was consistently identified across diagnosis, CR1, and REL1 disease stages. The majority of putative interactions identified at diagnosis or REL1 were unique to each disease stage, despite being sampled from the same patient cohort: 32/49 (65.3%) unique to diagnosis and 18/28 (64.3%) unique to REL1. One such interaction pair was chr8gain-*TET2*, which was only observed to be statistically significant at the time of diagnosis (**Supplementary Figure 4D-E**). Our analyses therefore suggest that many genetic interactions in AML exhibit a certain degree of context-dependence, demonstrating the importance of interrogating the genomic features of AML across diverse disease stages.

### Early DNMT3A and TET2 mutations differentially shape the evolutionary trajectories of AML

We next sought to determine how early CH mutations influence downstream mutation stability at CR1 and loss/gain at REL1. We categorized patients by their mutational status in *DNMT3A*, *TET2*, and *ASXL1* at the time of diagnosis (**Figure 4A**). Comparing pairs of DTA genes, we assessed cohort-level mutation frequencies in each of these patients at the time of diagnosis and relapse (**Supplemental Figure 5A-F**). While *FLT3* mutations were observed across patients with any of the DTA mutations, *FLT3* mutations were uniquely enriched in *DNMT3A*^mut^ cases at diagnosis (*p* = 0.01) and relapse (*p* = 0.02) (**Figure 5B**). Meanwhile, *NPM1* mutations were significantly enriched in *DNMT3A*^mut^ cases at both diagnosis (*p* = 9.8*10^-11^) and relapse (*p* = 5.5*10^-7^), as well as in *TET2*^mut^ cases – albeit to a lesser extent – both at diagnosis (*p* = 0.03) and relapse (*p* = 0.02). In contrast, *CBL* mutations were uniquely enriched in *TET2*^mut^ samples at all three stages of disease: diagnosis (*p* = 0.009), remission (*p* = 0.03) and relapse (*p* = 0.008). Notably, none of these mutations (*FLT3*, *NPM1*, and *CBL*) showed significant association with *ASXL1*^mut^ patients; rather these samples showed an enrichment for *SRSF2* mutations at diagnosis (*p* = 0.02) and remission (*p* = 0.01) (**Figure 4B**). Given the divergent genetic associations with distinct DTA mutations, and the relative enrichment of *CBL* and *SRSF2* mutations in myelodysplastic syndrome (MDS), we wondered if these findings generalized beyond our *de novo* AML cohort. We analyzed a cohort of untreated MDS patients^39^, and observed strong enrichment of *NPM1* mutations exclusively in *DNMT3A*^mut^ patients, while *CBL* and *SRSF2* mutations were associated with both *TET2* and *ASXL1* alterations (**Figure 4C**). *FLT3* mutations were observed at similar frequencies between *DNMT3A*^mut^ and *ASXL1*^mut^ samples. Collectively, these results indicate that early CH-related mutations show distinct mutational partners that persist at multiple stages of disease development and after therapy. While some of these co-mutational patterns are conserved between *de novo* AML and MDS, there are nevertheless important distinctions between the two, suggesting differences in the genetic interaction networks driving genomic evolution in these disease states.

**Figure 4:**
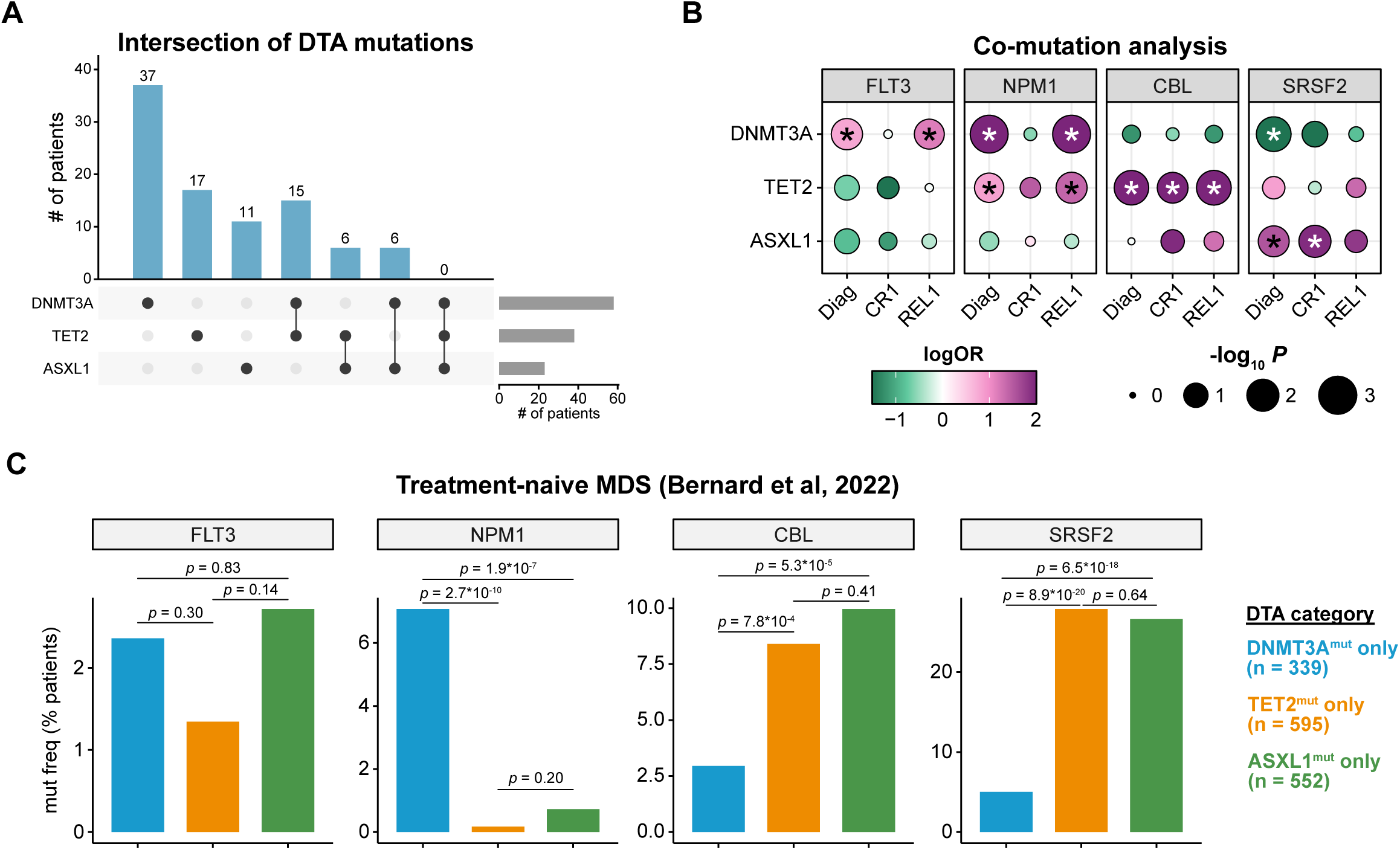
Early mutations in *DNMT3*A, *TET2*, and *ASXL1* differentially shape the subsequent evolution of AML from diagnosis through relapse. **A.** Upset plot indicating the number of patients at diagnosis with mutations in *DNMT3A, TET2* and *ASXL1.* The number of patients per group is indicated above each bar. **B.** Co-mutation analysis of *FLT3, NPM1, CBL*, and *SRSF2* in relation to *DNMT3A, TET2* or *ASXL1* across the entire cohort. Dots are color-coded by logORs and size-scaled by statistical significance (Fisher’s two-sided exact test). Asterisks denote *p* < 0.05. **C.** Bar plot detailing the frequency of *FLT3*, *NPM1*, *CBL*, or *SRSF2* mutations in a cohort of patients with untreated myelodysplastic syndrome (MDS)^39^, stratified by *DNMT3A*, *TET2*, and *ASXL1* mutation status. Statistical significance was assessed by Fisher’s two-sided exact test.

**Figure 5:**
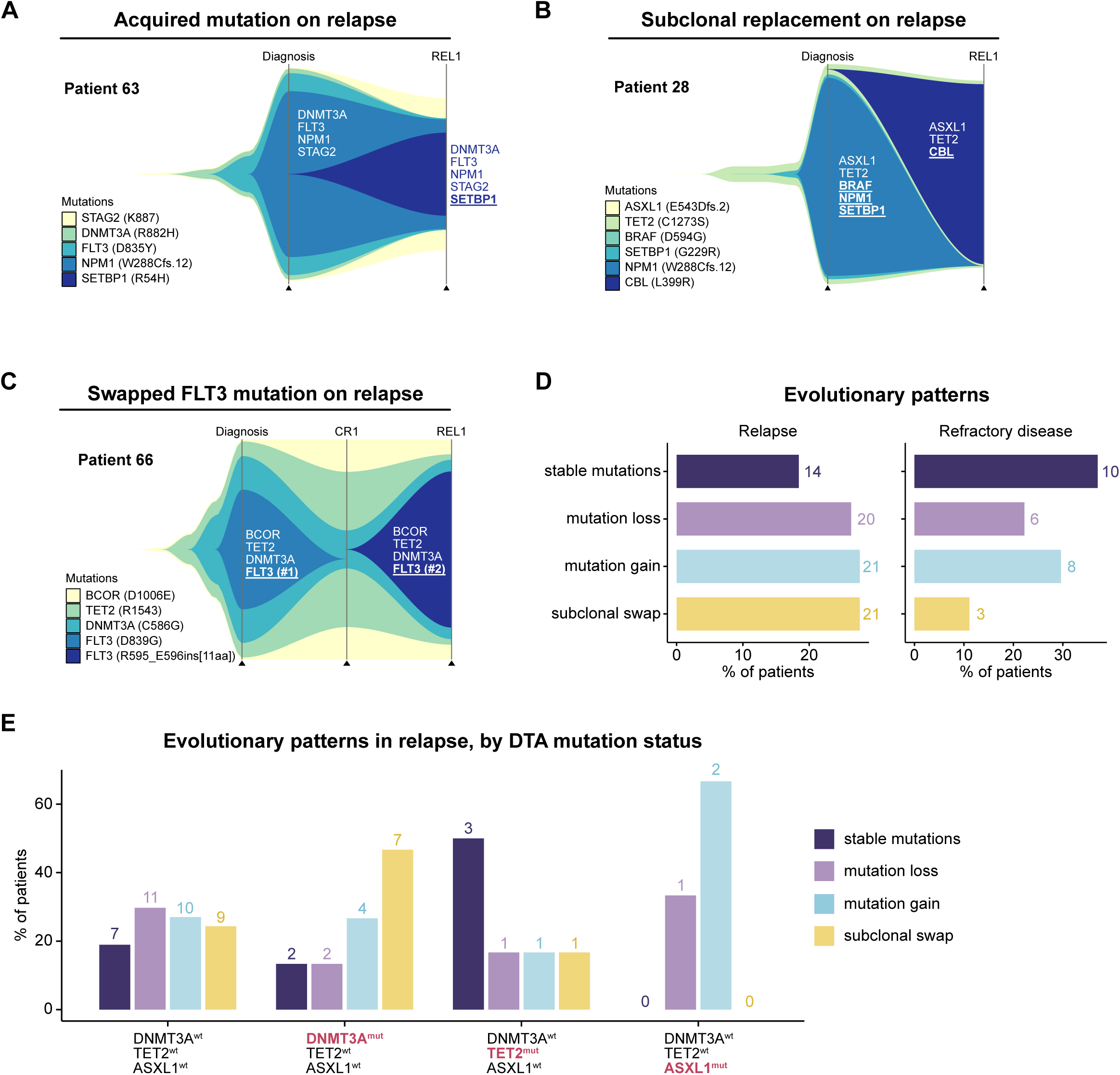
Patterns of AML genomic evolution from diagnosis to relapse. **A-C.** Fish plots detailing the expansion and contraction of specific variants within individual patients from diagnosis to relapse. **D.** Classification of evolutionary patterns at the time of relapse (left) or refractory disease (right) across the entire cohort. **E.** Bar plot detailing the type of relapse patterns observed in patients jointly stratified by *DNMT3A, TET2* and *ASXL1* mutation status.

### Patterns of AML genomic evolution from diagnosis to relapse

As the prior analyses were performed on the cohort-level, comparing mutation frequencies in different cross-sections of the AML disease course, we next sought to explore the characteristics of AML evolution within individual patients. We therefore applied the CALDER algorithm^40^ to help infer and visualize phylogenetic relationships from matched longitudinal AML sequencing data in individual patients.

Among the patterns of AML evolution from diagnosis to relapse, we observed several cases in which, at the time of relapse, an additional driver mutation had been acquired on top of the original mutations that were seen on diagnosis (**Figure 5A**). For instance, Patient 63 had a dominant clone at diagnosis with mutations in *FLT3*, *NPM1*, *DNMT3A*, and *STAG2*. At the time of relapse following induction chemotherapy, the above four mutations were still present, but now the cells had acquired an additional *SETBP1* mutation. In such patients, it is likely that the initial AML clone observed at diagnosis subsequently re-expanded following incomplete elimination by induction chemotherapy, gaining additional mutations in the process. In other cases, we observed evidence of subclonal replacement, with reciprocal loss and gain of driver mutations as a consequence of therapy and subsequent relapse (**Figure 5B**). As an example, Patient 28 had a dominant clone on diagnosis with *ASXL1*, *TET2*, *BRAF*, *NPM1*, and *SETBP1* mutations; on relapse, the dominant clone had lost the *BRAF, NPM1*, and *SETBP1* mutations, instead acquiring a *CBL* mutation. Among such cases of subclonal replacement, we observed cases in which a *FLT3* mutation seen at the time of diagnosis was subsequently replaced by a different *FLT3* alteration on relapse (**Figure 5C**). For Patient 66, the initial *FLT3*^D839G^ mutation was replaced with a *FLT3* internal tandem duplication (ITD) at relapse. Similarly, Patient 98 had a *FLT3* ITD mutation at diagnosis that was replaced with a distinct *FLT3* ITD mutation on relapse. It is likely that chemotherapy had successfully eliminated the AML-driving *FLT3*^mut^ clone, with subsequent disease relapse being driven by the acquisition or expansion of a distinct *FLT3*^mut^ clone. These cases demonstrate “convergent evolution” occurring within individual patients, consistent with the well-established role of *FLT3* mutations in driving malignant transformation.

We next analyzed the evolutionary trajectories for all patients profiled at diagnosis and relapse (n = 76) and classified them into one of four broad relapse patterns. Patients were distributed across these four categories, with 14/76 (18.4%) of cases demonstrating stable mutational profiles, 21/76 (27.6%) acquiring a new mutation, 20/76 (26.3%) losing an initial mutation, and 21/76 (27.6%) exhibiting subclonal swaps (**Figure 5D**). In patients sequenced at the time of refractory disease (n = 27), we observed that comparatively fewer patients (3/27, 11.1%) underwent subclonal swaps while stable mutational profiles were most common (10/27, 37%). Across all patients, subclonal swapping trended towards being more common in relapse compared to refractory disease (*p* = 0.11). When specifically comparing the relative proportions of stable mutational profiles to mutational swaps, mutational swaps were more common in relapse than in refractory disease (*p* = 0.049). Finally, we sought to determine how founding CH-mutations in *DNMT3A, TET2,* or *ASXL1* influenced evolutionary trajectories in relapse. While there were limited sample sizes for patients with only a single DTA mutation (*DNMT3A, TET2,* or *ASXL1*), we observed that *DNMT3A*^mut^ (*TET2*^wt^, *ASXL1*^wt^) AML was more likely to relapse through subclonal swaps (7/15, 46.7%) than *TET2*^mut^ (*DNMT3A*^wt^, *ASXL1*^wt^) AML (1/6, 16.7%). On the other hand, *TET2*^mut^ (*DNMT3A*^wt^, *ASXL1*^wt^) AML more often relapsed with stable mutation profiles (3/6, 50%) than *DNMT3A*^mut^ (*TET2*^wt^, *ASXL1*^wt^) AML (2/15, 16%) (**Figure 5F)**. Taken together, these analyses showcase the utility of longitudinal genomic profiling to reveal recurrent evolutionary modes of AML relapse across individual patients.

## DISCUSSION

While other large cohorts of patients have been analyzed with both exome sequencing and panel-based approaches, most include mixtures of AML evolved from a prior MDS, *de novo* AML, and relapsed/refractory AML. Our retrospective study specifically focused on patients with no prior hematological diagnoses or hematopoietic abnormalities. Similar studies have been performed retrospectively on clinical trial samples^27^, including those specifically focused on *FLT3* mutant^24,25^ or *NPM1* mutant^22,41^ AML. These studies were largely executed in the research setting using exome wide assays.^42^ Both a strength and limitation of our study was the use of a CLIA-approved targeted gene panel; as our study is built on real-world data collected as part of routine clinical practice, our findings are directly relevant to clinicians and patients. However, the technical limitations of our NGS panel likely leads to underestimation of mutation evolutionary processes, as we did not query genes outside of the panel. The NGS panel at our institution was also updated throughout the course of the study, with subsequent versions including additional genes; in the current study, however, we did not identify any variants that were exclusively detected at later timepoints solely due to discordant panel versions. Another important limitation is that our clinical sequencing and analysis pipeline allowed for a minimum 2-4% VAF cutoff for reporting variants. Error corrected sequencing approaches have demonstrated that VAFs as low as 10^-5^ can offer prognostic information for *FLT3* and *NPM1* mutations in the context of measurable residual disease detection at first remission.^36,37^ The paucity of *NPM1* mutations detected at CR1, and their near uniform recurrence at relapse, suggests that our dataset is likely enriched for false negatives for *NPM1,* and potentially other genes, at remission. While these limitations are important to recognize, our study represents real-world data presented to clinicians at the time of diagnosis, remission and relapse, using standard sequencing approaches in routine clinical practice.

To our knowledge, no study to date has systematically compared the genomic profiles of *DNMT3A*^mut^ and *TET2*^mut^ AML as the disease evolves from initial diagnosis through remission and subsequent relapse. *DNMT3A* and *TET2* are the most commonly mutated genes associated with CH^7–9^, and both of these genes encode key regulators of DNA methylation.^43^ As *DNMT3A* and *TET2* mutations are among the earliest genetic alterations in AML, dysregulation of DNA methylation is presumably an important predisposing factor for the subsequent pathogenesis of AML. Curiously, however, DNMT3A and TET2 play diametrically opposing roles in DNA methylation: whereas DNMT3A catalyzes DNA methylation, TET2 demethylates DNA. It stands to reason, then, that the evolutionary fitness landscapes of malignancies arising from *DNMT3A*^mut^ clones likely differ from those that derive from *TET2*^mut^ clones. Our analyses illuminate the distinct genomic features of *DNMT3A*^mut^, *TET2*^mut^, and *ASXL1*^mut^ AML. While *DNMT3A*^mut^ AML is comparatively enriched in *FLT3* and *NPM1* mutations^4^, *TET2*^mut^ and *ASXL1*^mut^ AML are instead enriched in *CBL* and *SRSF2* mutations. We further demonstrate that these differences persist through chemotherapy and are often conserved at diagnosis and relapse. As *DNMT3A*, *TET2*, and *ASXL1* mutations represent the earliest genetic events in the pathogenesis of AML, our findings demonstrate how “founding” preleukemic driver mutations can subsequently mold the evolutionary paths traversed in the course of AML evolution.

Of note, we had carefully curated the present cohort to exclude patients that had preexisting MDS prior to AML diagnosis. As the co-occurring module of *TET2*, *CBL*, and *SRSF2* mutations is highly prevalent in MDS^39^, it is interesting that we were able to recapitulate this mutational pattern in our cohort of *de novo* AML. We further found that the co-occurring module of *DNMT3A* and *NPM1* mutations that we observed in *de novo* AML was also seen in the MDS cohort. While there were distinctions between the mutational archetypes seen in each cohort, these commonalities suggest that regardless of whether AML arises *de novo* or as a gradual progression from MDS, the underlying genetic interactions shaping their evolutionary trajectories appear to be broadly conserved. These data are consistent with the ELN2022 guidelines of *NPM1* mutations being sufficient to diagnose patients with AML, that might otherwise fit histopathological descriptions of MDS.^44^ Our data support the notion that *DNMT3A*^mut^ MDS likely encompasses a genomic co-mutational landscape that is reminiscent of co-mutational partners found in AML.

Moving forward, further mechanistic studies are needed to understand the molecular basis underlying the divergent mutational trajectories of *DNMT3A*^mut^ vs *TET2*^mut^ AML. Given their opposing functions in DNA methylation, we anticipate that the distinct epigenetic changes associated with *DNMT3A* vs *TET2* loss-of-function act to differentially pre-pattern the epigenetic landscape on which hematopoietic stem/progenitor cells subsequently evolve into AML.^45^ We speculate that these early epigenetic differences can impact the fitness effects of subsequent AML driver mutations, leading to divergent evolutionary trajectories through diagnosis and relapse.

## Methods

All sequencing results, ancillary studies, and clinical information were collected retrospectively in accordance with protocols approved by the Institutional Review Board at the University of Pennsylvania.

### Patient selection and data collection

We searched internal pathology databases for all patients with two or more NGS studies performed at least 30 days apart on blood or bone marrow specimens between February 14, 2013 and June 31, 2018 using our institution’s clinical targeted hematologic malignancies NGS panel. Patients with testing performed at initial diagnosis of *de novo* AML and subsequent testing performed at cytologic complete remission (CR), relapse (REL), or disease refractory to initial therapy for de novo AML (REF) were included. Remission, relapse, and refractory states were determined from review of clinical notes from the electronic medical record (EMR) and corresponding hematopathology studies performed on bone marrow specimens. CR was defined as having morphologic evidence on bone marrow biopsy of trilineage hematopoiesis and <5% blasts. Subjects with diagnoses of therapy-related AML or AML with myelodysplasia-related changes were excluded. Cytogenetics, treatment history, and demographic details were also retrospectively recorded from the electronic medical record (Supplementary Table 1).

### Genomic sequencing

All patients were sequenced at the same institution on a clinically validated and CLIA-certified customized NGS panel which covers targeted coding regions and splicing junctions of genes that are commonly mutated in myeloid malignancies. All tests were ordered by treating physicians for clinical purposes.

DNA was extracted from fresh bone marrow aspirate or whole blood samples, and targeted sequencing of hot spots in exomes of 33 genes (HemeV1 panel, 2/14/2013 to 4/21/2015) or 68 genes (HemeV2 panel, 4/22/2015 to present) was performed using an Illumina TruSeq Custom Amplicon assay that was optimized to identify mutations with known or suspected associations in the pathogenesis of myeloid malignancies, as well as some mutations enriched in a subset of lymphoid neoplasms. Matched-normal samples were not submitted for any of the patients.

The first version of the panel included hotspots from the following genes (350 total amplicons): *ASXL1, ATM, BRAF, CBL, CDKN2A, DDX3X, DNMT3A, ETV6, EZH2, FBXW7, FLT3, GNAS, IDH1, IDH2, JAK2, KIT, KLHL6, KRAS, MAPK1, MYD88, NOTCH1, NPM1, NRAS, PHF6, PTEN, PTPN11, RUNX1, SF3B1, TET2, TP53, WT1, XPO1, ZMYM3*. The second version of the panel added hotspots from the following genes (673 total amplicons): *ABL1, BCOR, BCORL1, BIRC3, CALR, CEBPA, CSF1R, CSF3R, BRINP3 (FAM5C), GATA2, HNRNPK, IL7R, MAP2K1, MIR142, MPL, MYC, MYCN, NF1, NOTCH2, PDGFRA, POT1, PRPF40B, RAD21, RIT1, SETBP1, SF1, SF3A1, SMC1A, SRSF2, STAG2, TBL1XR1, TPMT, U2AF1, U2AF2, and ZRSR2*. As *CEBPA* testing is performed only upon request, particularly at diagnosis, this data was excluded from the analysis as not all patients were routinely tested for it.

### Variant calling and annotation

NGS data was processed through a custom in-house bioinformatics pipeline that was clinically validated to call single nucleotide variants (SNVs) at a frequency of 2-4% and small insertions and deletions (indels) at a frequency of 1%. The minimum mean coverage was 2500x across the entire panel and the minimum read depth for each amplicon was 250x. The lowest reportable variant allele frequency was 2% for SNVs in *FLT3* and *NPM1* and 4% for mutations in all other genes in the panel. Variants passing these filtering criteria were included for analysis, regardless of pathogenicity classifications.

### Data analysis

For simplicity, if multiple NGS studies were conducted at the same “stage” of disease (eg CR1, REL1), only the earliest NGS sample was retained for further analysis. To compare the mutation frequencies of genes between different groups, we used Fisher’s two-sided exact test. We classified variants as exclusive or shared across disease stages using a binary classification schema (i.e., present or absent), based on the variants that were reported following the variant calling approach described above. To calculate ΔVAFs, we directly subtracted the VAFs between disease stages, taking care to match the same variants within individual patients by the annotated amino acid changes.

For genetic interaction analyses, we compared the co-mutation frequencies for each gene pair using Fisher’s two-sided exact test. For visual clarity in the figures, we omitted gene pairs that did not meet the nominal significance threshold of *p* < 0.05. Data were reported in terms of log odds ratios (ORs). For analysis of mutational co-occurrence patterns in the MDS cohort^39^, we extracted data using the cBioPortal browser^46,47^ and used Fisher’s two-sided exact test.

To construct parsimonious phylogenies for the longitudinal NGS data, we used the CALDER algorithm^40^. For each patient, we included all identified variants (expressed in terms of VAFs) at diagnosis, CR1, and/or REL1. We visualized the resulting phylogenies using clevRvis.^48^ To classify relapse patterns into different categories, we manually reviewed the longitudinal changes in VAFs within each patient. If all mutations observed at diagnosis were again observed at REL1 with no additional or lost mutations, the relapse pattern was classified as “stable mutations.” Accordingly, if the REL1 mutation profile was the same as the diagnosis mutation profile, but with the addition or loss of one or more mutations, these cases were classified as “mutation gain” or “mutation loss.” If the REL1 sample had acquired one or more new mutations while also losing one or more mutations that were originally present at diagnosis, we classified it as a “subclonal swap”.

To compare the mutation frequencies observed in our cohort at diagnosis to previously published datasets (TCGA-AML^4^ and OHSU-AML^38^), we extracted data using the cBioPortal browser.^46,47^ We included all genes that were mutated in ≥ 4% of our cohort for analysis. To compare mutation frequencies between cohorts, we calculated Spearman and Pearson correlation statistics.

## Data availability

All mutation calls and clinical annotations are publicly available on Github: https://github.com/rdchow/PennAML.

## Code availability

All analysis code is publicly available on Github: https://github.com/rdchow/PennAML.

## Supplementary Tables

**Table S1:** All called mutations and karyotype aberrations across all samples.

**Table S2:** Filtered mutations and karyotype aberrations in the final analysis set.

**Table S3:** Clinical annotations of all samples.

**Table S4:** Filtered clinical annotations of samples included in the final analysis set.

**Table S5:** Comparison of mutation frequencies for each gene across disease stages.

**Table S6:** Longitudinal tracking of variant allele frequencies in individual patients.

**Table S7:** Co-mutation analysis at the time of diagnosis.

**Table S8:** Co-mutation analysis at the time of first remission.

**Table S9:** Co-mutation analysis at the time of first relapse.

**Table S10:** Patterns of genomic evolution

## ACKNOWLEDGEMENTS

R.L.B. was supported by the National Cancer Institute (R00CA248460, UG1CA233332), American Society of Hematology and the Leukemia Research Foundation.

## AUTHOR CONTRIBUTIONS

P.V., and J.M. conceived and designed the study. R.D.C. designed and executed the experimental analysis. R.D.C., P.V., S.D., J.M., A.Y., and N.S. performed experimental analysis and curated patient records and data. J.M. and R.L.B., supervised the study. R.D.C. and R.L.B. wrote the manuscript with significant revisions and critical feedback from P.V., S.M.L., and J.M.; all authors reviewed and commented on the final manuscript.

## Supplementary Figure Legends

**Supplementary Figure 1:**
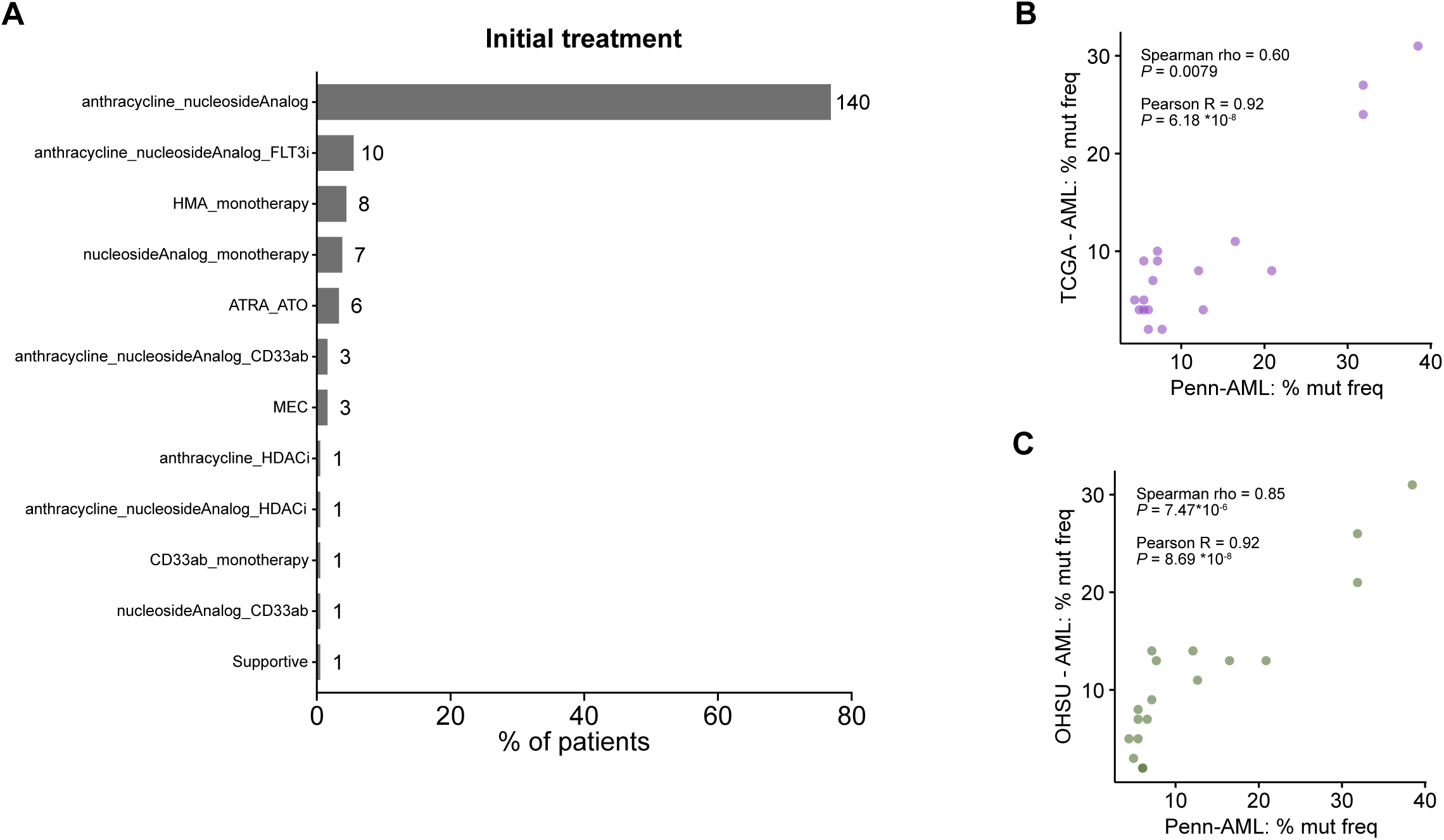
Comparison of mutation frequencies across AML patient cohorts. **A.** Bar plot depicting distribution of initial treatments in the Penn AML cohort. **B-C.** Comparison of gene mutation frequencies in the current cohort (Penn-AML) vs (**B**) TCGA-AML or (**C**) BeatAML. The associated Spearman and Pearson correlation statistics are annotated. Genes were filtered to those that were mutated in ≥ 4% of patients in the Penn-AML cohort at diagnosis.

**Supplementary Figure 2:**
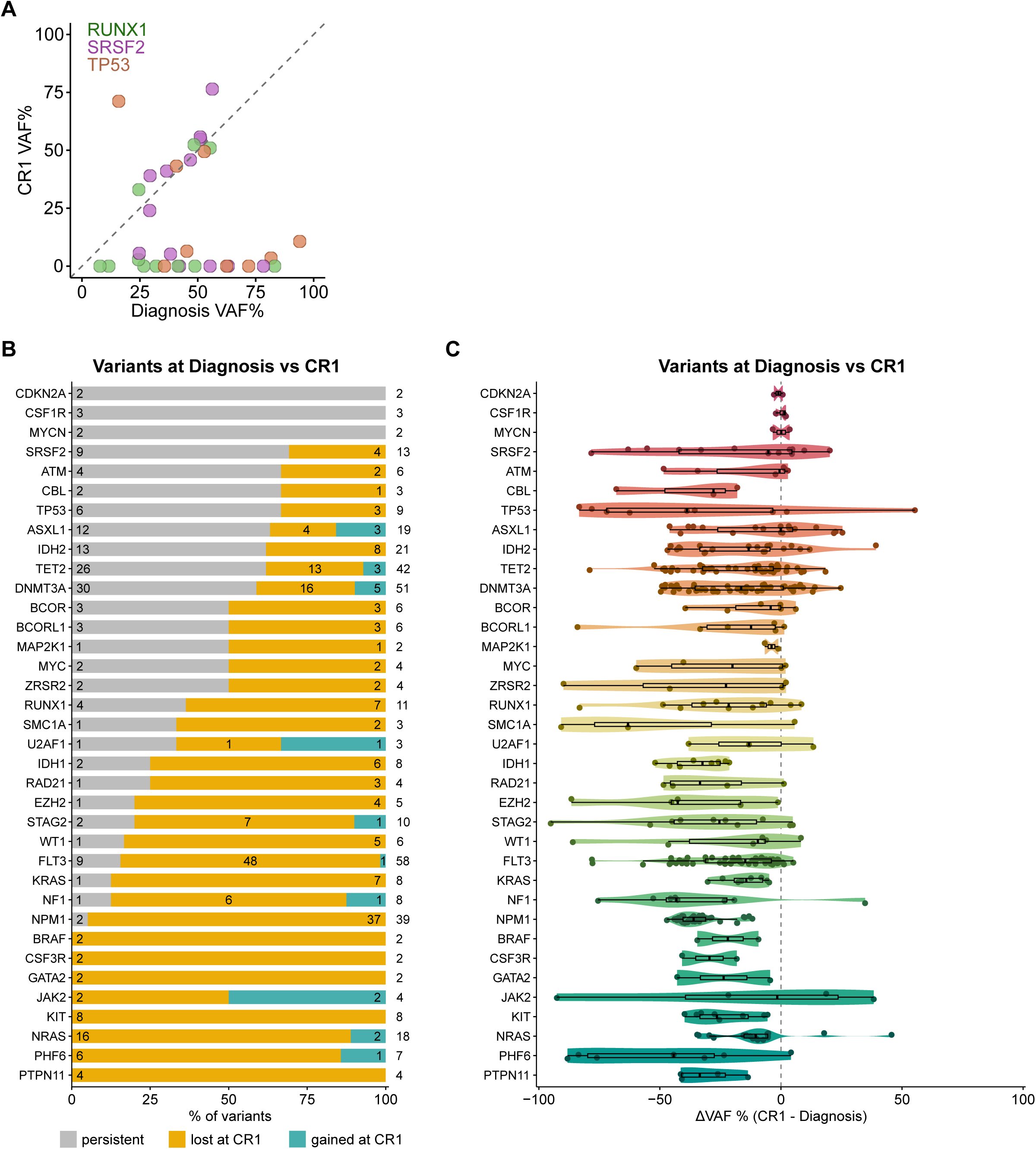
Cohort-wide patterns of mutations gained and lost at first remission. **A.** Comparison of VAFs at diagnosis vs CR1 for *RUNX1*, *SRSF2* and *TP53*. Each point represents one variant in a specific patient across time. Dashed lines denote equality between disease stages. **B.** Bar plot depicting the relative proportions of different mutation trajectories between diagnosis and CR1 in individual patients, filtered for genes with at least two variants identified across the cohort. Colors indicate whether the mutation was stable (grey), lost (yellow) or gained (teal) from diagnosis through CR1. The number to the right indicates the total number of variants identified among paired diagnosis and remission samples for the indicated gene; within each section of the bar plot, the numbers indicate the number of variants within each category. **C.** Violin plot depicting the difference in VAFs (ΔVAF) between CR1 and diagnosis. Negative values indicate a lower VAF at CR1, while positive values indicate a higher VAF.

**Supplementary Figure 3:**
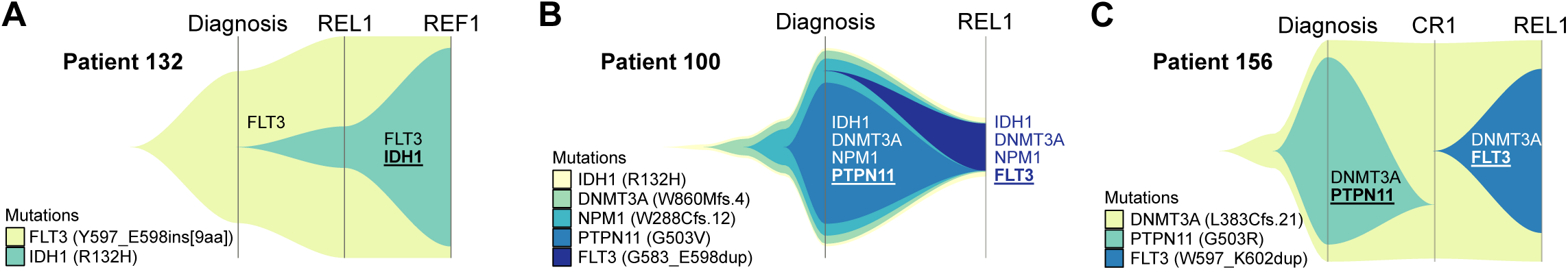
Representative examples of observed mutational patterns. **A-C.** Fishplots detailing the expansion and contraction of specific variants within individual patients from diagnosis to relapse. **A. C**lonal evolution of patient 132, showing the acquisition of an *IDH1* mutation that subsequently expanded through refractory disease. **B-C.** Clonal evolution in patients 100 and 156, where an initial *PTPN11*^mut^ clone was eliminated by treatment and subsequently replaced with a *FLT3*^mut^ clone.

**Supplementary Figure 4:**
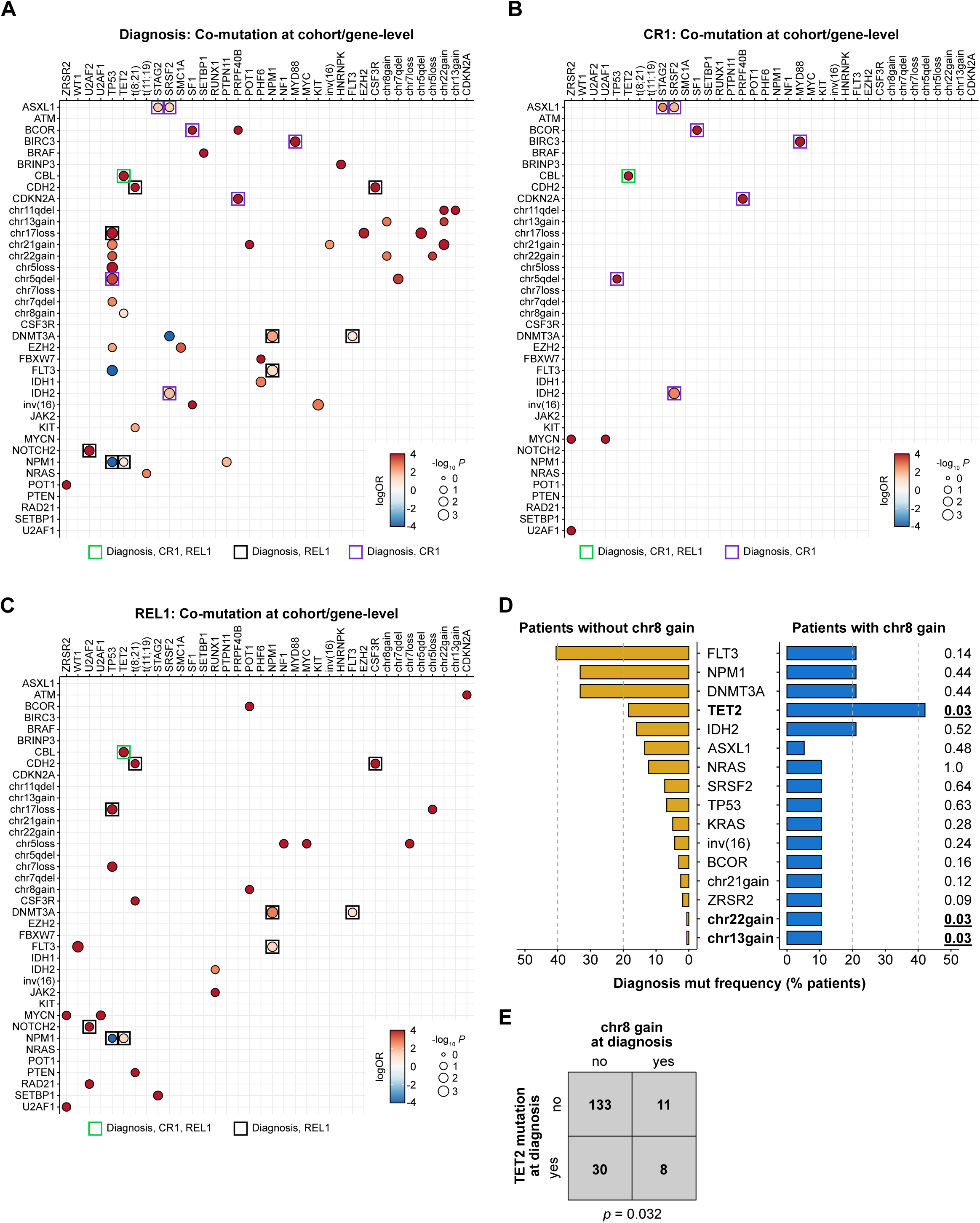
Conserved and disease stage-specific genetic interactions in AML A-C. Pairwise co-occurrence analysis for mutations or karyotype aberrations, **(A)** at diagnosis, **(B)** at CR1, and **(C)** at REL1. Visualized alterations were filtered to those comprising a significant genetic interaction at diagnosis and/or at REL1. Dots are color-coded by logORs and size-scaled by statistical significance (Fisher’s two-sided exact test). Alteration pairs that were not statistically significant (*p* ≥ 0.05) are omitted for clarity. Alteration pairs that were significant both at diagnosis and REL1 are highlighted with a black box, while those significant at diagnosis and CR1 are highlighted in a purple box. Alteration pairs that were significant across diagnosis, CR1, and REL1 are outlined in green. **D.** Mutation frequencies of select genomic alterations at time of diagnosis in patients with (right) or without (left) chr8 gain. Alterations with ≥ 10% mutation frequency in patients either with or without chr8 gain are shown. The associated *p*-values by Fisher’s two-sided exact test are shown on the far right. **E.** Co-occurrence matrix of chr8 gain and *TET2* mutation at time of diagnosis, assessed by Fisher’s two-sided exact test.

**Supplementary Figure 5:**
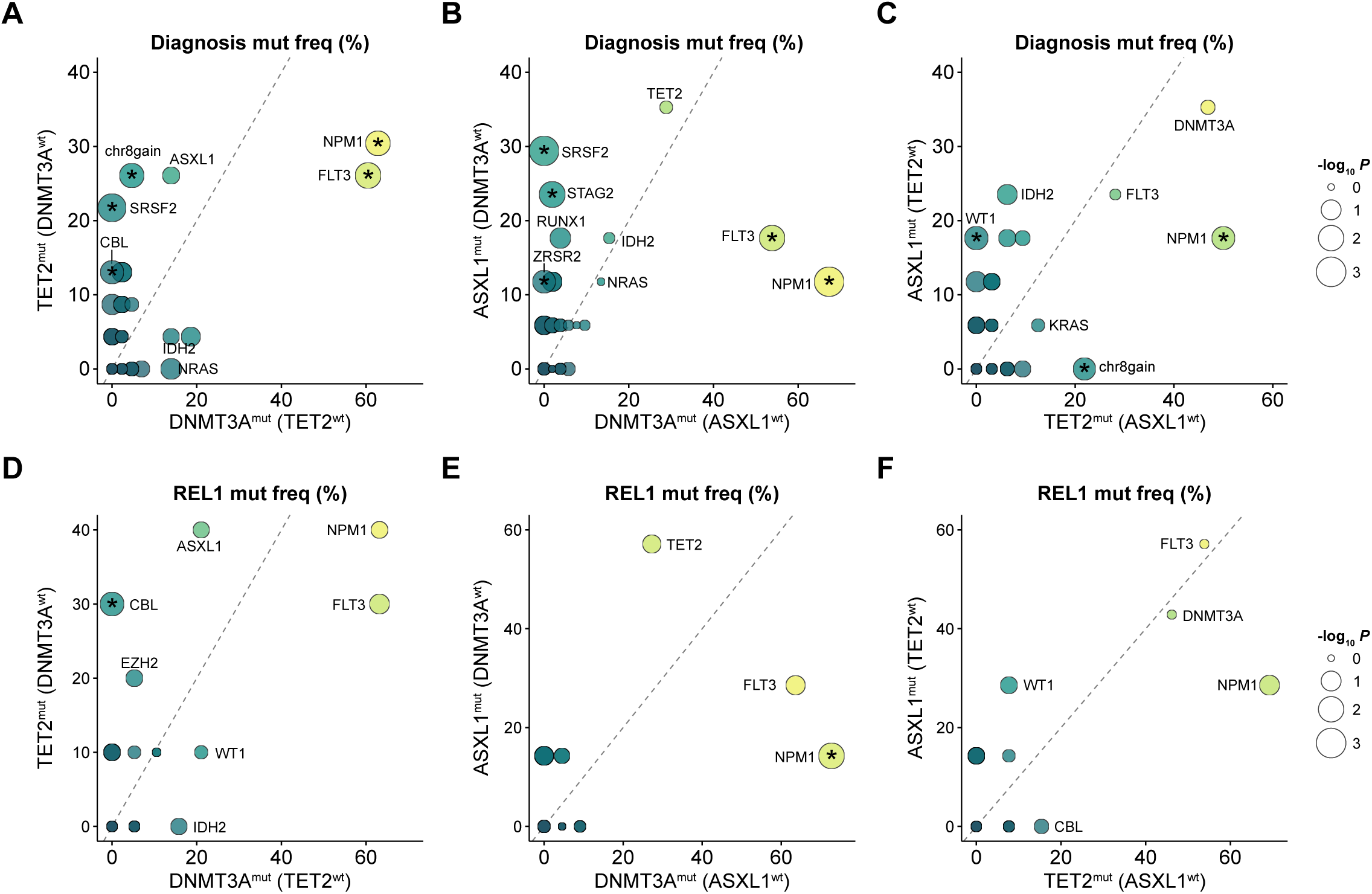
Comparison of co-mutational frequencies for distinct DTA mutant groups at diagnosis and relapse. **A-F.** Direct comparison of mutation frequencies between *DNMT3A*^mut^ vs *TET2*^mut^ (**A,D**), *DNMT3A*^mut^ vs *ASXL1*^mut^ (**B,E**) and *TET2*^mut^ vs *ASXL1*^mut^ (**C,F**) samples at the time of diagnosis (**A-C**) or REL1 (**D-F**). For relapse samples, the classification of *DNMT3A*^mut^, *TET2*^mut^ and *ASXL1*^mut^ was based on their mutational profile at time of diagnosis. Point sizes are scaled by statistical significance (Fisher’s two-sided exact test) and colored based on mutation frequency. Genes reaching the statistical significance threshold are denoted with an asterisk. Dashed lines denote equality between the two categories.

